# Climate influences scrub typhus occurrence in Vellore, Tamil Nadu, India: Analysis of a 15 year dataset

**DOI:** 10.1101/2023.07.20.23292936

**Authors:** Solomon D’Cruz, Kotamreddy Sreedevi, Cheryl Lynette, Karthik Gunasekaran, JAJ Prakash

## Abstract

**Background:** Climate is one of the major factors determining the prevalence and seasonality of vector borne diseases like scrub typhus (ST). We analyzed, the association of the meteorological factors like temperature, rainfall and humidity with scrub typhus using the 15 years scrub typhus data from a tertiary care hospital in Vellore, South India.

**Methods:** Demographic data of permanent residents of Vellore, who had IgM ELISA results for scrub typhus for the time period of May 2005 to April 2020were included. Meteorological data was correlated with the monthly scrub typhus cases; negative binomial regression model was used to predict the relation between scrub typhus occurrence and climate factors.

**Results:** Maximum number of ST cases were reported between the months August and February with October recording the highest number of cases. Elderly people, farmers, agricultural workers and housewives were associated with ST positivity significantly. For an increase of 1°C in mean temperature, the monthly ST cases reduced by 18.78% (95% CI: −24.12, −13.15%). On the contrary, for 1 percent increase in mean relative humidity (RH), there is an increase of 7.57% (95% CI: 5.44, 9.86%) of monthly ST cases. Similarly, an increase of 1mm of rainfall contributed to 0.50 to 0.70% of monthly ST cases (after two months) depending on the variables included in the analysis.

**Conclusion:** This study provides further information on the role of rainfall, temperature and humidity on the seasonality of scrub typhus in Vellore, South India. This baseline data will be useful for further analysis using spatio-temporal tools to better understand the seasonality in other parts of India.

## Introduction

Scrub typhus is caused by the obligate intracellular bacteria *Orientia tsutsugamushi*(1,2). The infection is transmitted to humans through the bite of larvae (chiggers) of trombiculid mites which act both as the vector and reservoir of the infection (3) and are common ecto-parasites of rodents (4). The chiggers get attached to many species of small rodents, particularly wild rats of the subgenus *Rattus* which is a natural host for *Orientia tsutsugamushi* (5). The chigger infection with scrub typhus is perpetuated and maintained in nature by transovarial and transstadial transmission (6).

Humans are the accidental host for the infection as they can pick up the mite when they walk, sit or lie down in the ground infested by chigger mites (5). A necrotic lesion, eschar which appears at the site of chigger bite - the mite bite site (7,8). Once infected, the patients may have symptoms like fever, headache, myalgia, cough, generalized lymphadenopathy, nausea, vomiting and abdominal pain. Of these, fever and head-ache are the most commonly reported symptoms (1). Scrub typhus may also lead to severe complications like multi-organ dysfunction (MODS) and the resulting mortality can be very high among untreated cases (1,9).Prevalence of eschar among scrub positives ranges from 8% to 58% in Indian studies (10–14) and the reason for this less detection of eschar which is a black scab with erythematous halo is due to its painless nature occurring mostly in flexural areas and hence often overlooked (15). Diagnosing scrub typhus is mostly by clinical suspicion and serological test (16). Without the presence of eschar, the diagnosis of scrub typhus is challenging and often other causes of fever get explored (17). Once suspected, oral doxycycline for mild infections (1) and a combination of parenteral doxycycline and azithromycin for severe infections have been shown to be effective against scrub typhus (18).

The occurrence of ST is seasonal in the endemic areas (5) and in Vellore, the cooler months were noticed to have increased number of cases (19). This is probably due to more chiggers getting attached to the rodents (20). It has also been noted that scrub typhus incidence is associated with relative humidity, temperature and rainfall (8,9). Further, there can be country wide variation across seasons and in different geographical condition as observed in China (21). Therefore, we analyzed a 15 year dataset of scrub typhus and compared the role of climatic factors like rainfall, temperature and humidity on seasonality of scrub typhus in Vellore.

## Materials and Methods

### Ethical statement

No human subjects were directly involved in the study. The scrub typhus data was obtained from the electronic laboratory register of the Immunology Laboratories, Department of Clinical Microbiology, Christian Medical College, Vellore. All the patient identifying variables in the data were removed before the analysis. The approval to conduct this study was obtained from Institutional Review Board (IRB) and Ethics Committee (EC) of the Christian Medical College, Vellore vide IRB Min no. 9866 dated 20^th^ January 2016.

### Study Area

Vellore district in Tamil Nadu has an area of 6075 Sq.km (22). According to the census of India 2011, The population of Vellore was 39,36,331 with a population density of 648/sq. km(22). Vellore is bounded by Chittoor district of Andhra Pradesh in the North, Tiruvannamalai district in the South, Kancheepuram and Tiruvallur districts in the East and by Yelagiri hills in the West (Vellore District Statistical handbook 2016 – 17). It has 1,62,286 hectares of Forest area and the district is mostly surrounded by hills. The majority of the rainfall in this area is from the southwest monsoon. The major occupation of people in Vellore district is agriculture with paddy being the major crop grown. It has a forest cover of 27.94% with tropical climate (23).

**Figure 1:**
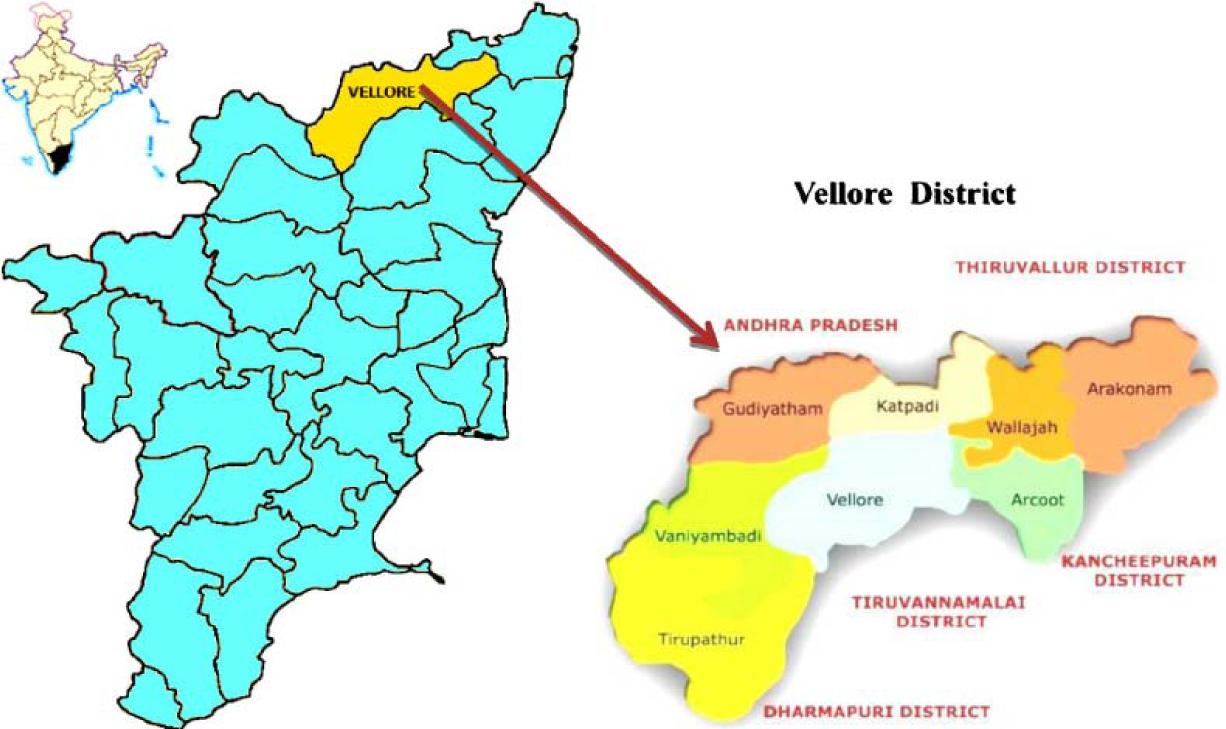
Map of Vellore district. Source:https://www.researchgate.net/figure/Map-showing-the-study-area-of-Vellore-India_fig1_282975836

### Methodology

The data for this research was obtained from the electronic medical records of a tertiary care hospital (Christian Medical College, Vellore), South India. The data included is for patients with suspected scrub typhus for whom the Scrub typhus IgM ELISA was requested. Data of such patients who underwent scrub typhus testing was obtained for the time period of May 2005 to April 2020. The diagnosis of scrub typhus was done based on the positivity of the serological test for scrub typhus (IgM), on individuals with acute undifferentiated febrile illness (AUFI), who were malarial parasites and blood culture negative. All the assays were performed with appropriate controls in the Immunology Laboratory (an ISO15189:2012 accredited lab) on an automated ELISA workstation (Euroimmun Analyzer I, Euroimmun AG, Lubeck, Germany). The cut-off OD value was set at 1.0 for scrub typhus positivity. Patients with OD values 1.0 and above in the IgM ELISA test were considered positive and those who were negative (OD <1.0) were used as a comparator group. Patients who are non-residents of Vellore district were excluded from the study.

Variables like age, sex, address, OP/IP, occupation, date of testing and results were obtained from the patient medical records of Christian Medical College, Vellore. Climatic factors like monthly mean maximum temperature, monthly mean minimum temperature, monthly mean relative maximum humidity, monthly mean relative minimum humidity and monthly total rainfall of Vellore was obtained from the Regional Meteorological Centre, Chennai.

### Statistical analysis

The data was entered in Microsoft excel spreadsheets (excel version 1997) and analyzed using IBM SPSS Statistics for Windows, Version 21.0 (IBM Corp, Armonk, NY, USA). The continuous variables were expressed in mean and standard deviation (SD) for normal distribution and median and inter-quartile range for skewed distribution. The categorical variables were expressed in frequencies and percentages. Spearman’s correlation (for skewed data) was used to correlate two continuous variables and 95% CI was reported. Mean temperature and mean humidity was calculated as monthly averages, whereas rainfall and ST cases were calculated as monthly aggregates. A negative binomial regression model was used to explore the relationship between monthly scrub typhus cases and meteorological factors. Since the scrub typhus data and rainfall data were overly dispersed we chose a negative binomial regression over the Poisson model. Spearman correlation coefficient matrix was done primarily to check for any existing correlation between the meteorological factors. This revealed a strong correlation of −0.739 (p <0.001) between mean temperature and mean humidity. To avoid collinearity problems, two negative binomial regression models were done namely model A and model B. Model A was computed using rainfall and mean temperature and model B was computed using rainfall and mean humidity. We calculated the influences (e^β^ – 1)* 100, which roughly corresponds to the percent increase, to quantify the effects of meteorological variables (24). The association between two categorical variables was analyzed using chi-square and logistic regression was done for adjusting the confounders.

## Results

From May 2005 to April 2020, 11001 were tested for scrub typhus, out of which 2784 were positives. The overall positivity rate among the people tested for ST is 25.3 percent. The male to female ratio is 1:1.4. The mean age of scrub typhus positive patients is 44.2 with a standard deviation of 19.2 (Median: 45, IQR: 30, 59).

**Table 1:** Demographic Characters.

| Demographic characters<br>(n = 11001) | Total tested | Negative | Positive |  | Positivity rate<br>(Positive/Total tested) |
| --- | --- | --- | --- | --- | --- |
|  |  |  | Numbers | Percentages |  |
| Sex |  |  |  |  |  |
| Male | 5570 | 4386 | 1184 | 42.5% | 21.3% |
| Female | 5431 | 3831 | 1600 | 57.5% | 29.5% |
| Total | 11001 | 8217 | 2784 | 100% | 25.3% |
| Age group |  |  |  |  |  |
| 0 - 20 years | 2036 | 1714 | 322 | 11.6% | 15.8% |
| 21 - 30 years | 2144 | 1745 | 399 | 14.3% | 18.6% |
| 31 - 40 years | 1699 | 1254 | 445 | 16% | 26.2% |
| 41 - 50 years | 1643 | 1144 | 499 | 17.9% | 30.4% |
| 51 - 60 years | 1588 | 1078 | 510 | 18.3% | 32.1% |
| >60 years | 1891 | 1282 | 609 | 21.9% | 32.2% |
| Total | 11001 | 8217 | 2784 | 100% | 25.3% |
| OP/IP |  |  |  |  |  |
| IP | 6902 | 5031 | 1871 | 67.2% | 27.1% |
| OP | 4020 | 3186 | 834 | 30% | 20.8% |
| Missing | 79 | 0 | 79 | 2.8% | NA |
| Total | 11001 | 8217 | 2784 | 100% | 25.3% |

**Figure 2:**
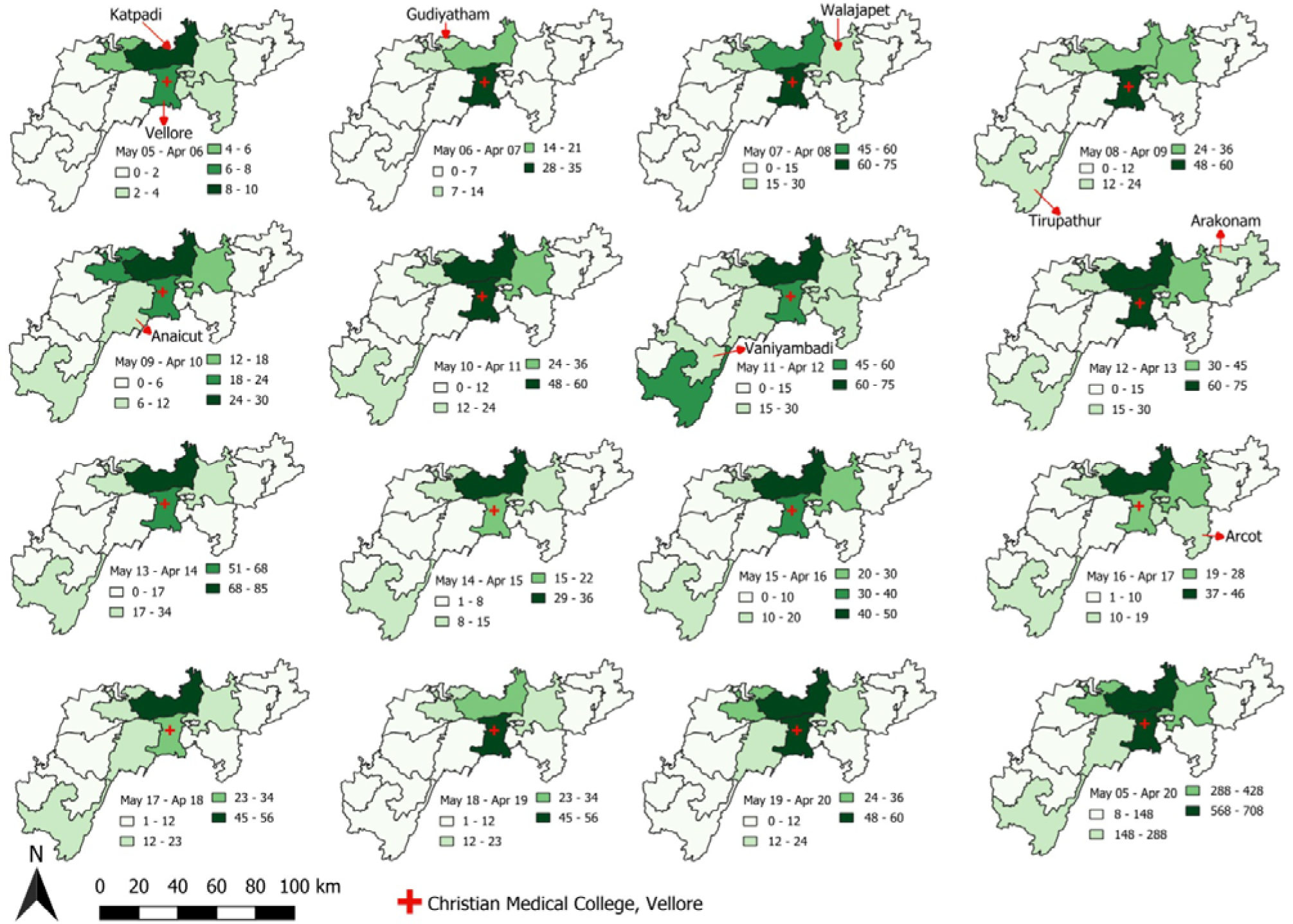
Spatiotemporal distribution of scrub typhus cases block wise in Vellore district from May 2005 – April 2020.

### Spatial and Temporal distributions

The average number of scrub typhus cases reported per year from the entire Vellore district is 186 with the majority of the cases from Katpadi and Vellore blocks with 45 and 44 cases per year respectively. Except for the year May 05 – April 06, Gudiyatham, Katpadi, Vellore and Walajapet blocks have contributed more than 10 scrub typhus cases in all the years.

The mean distance from the patient’s house to CMC hospital ranges from 18.1 km (SD: 19.8 km) in May 19 – April 20 to 31.4 km (SD: 26.5 km) in May 11 – April 12. The overall mean distance in all 15 years from the hospital to scrub typhus patients is 24.8 km (SD: 22.7 km) (Supplementary table 3).

**Figure 3:**
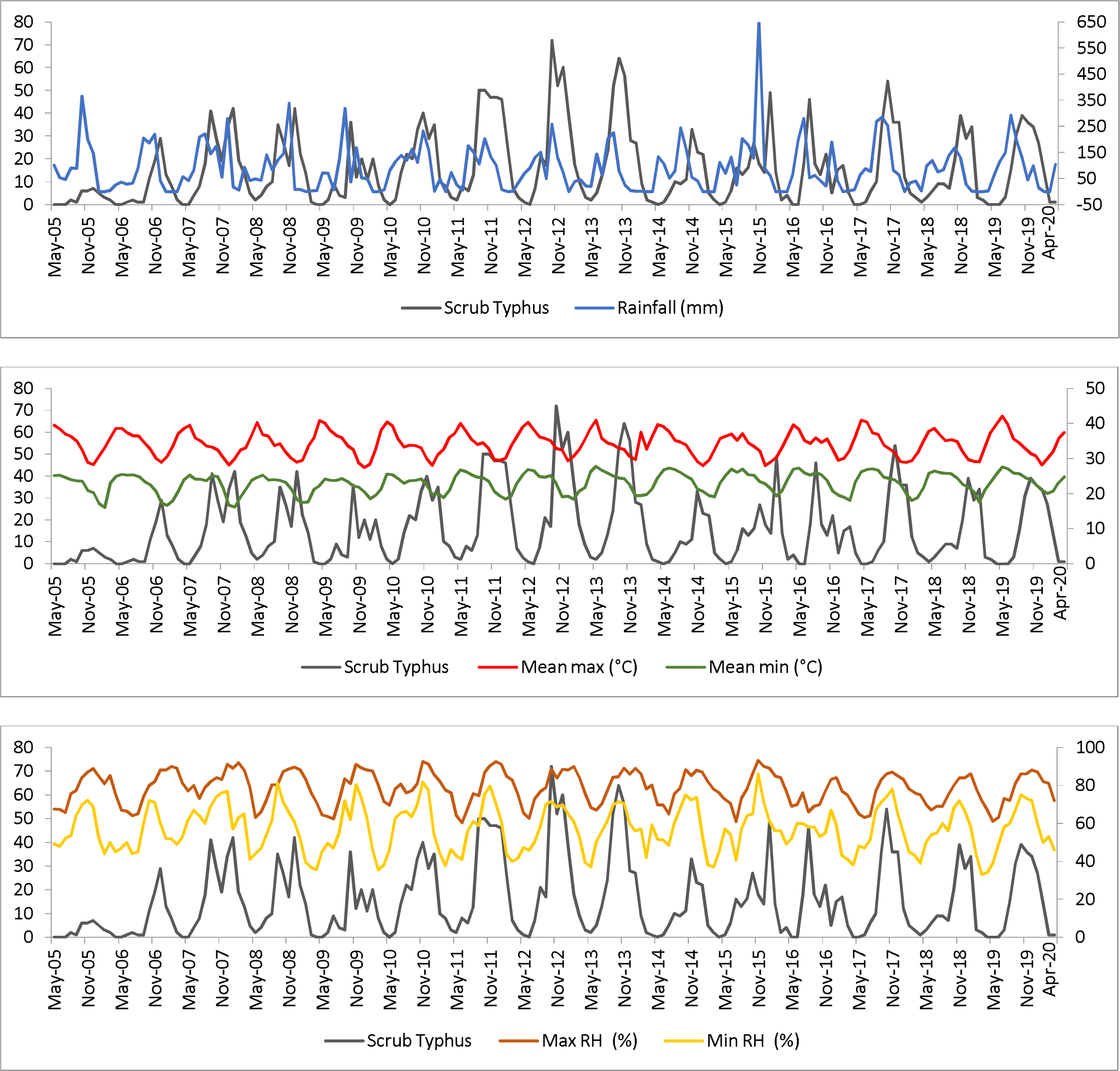
Month-wise trend of ST positives vs Rainfall, Temperature and Humidity from May 2005 – April 2020.

The above figure shows the month-wise trend of scrub typhus positive and the rainfall received, average temperature and humidity of that corresponding month. Increase in cases has occurred only after 2 months of the increased rainfall and it is uniformly observed in all 15 years. The number of ST cases gradually increased with the decrease of the temperature (Max and min) and decreased as the temperature increased (Max and min). This pattern is observed in all 15 years. With relative humidity, the ST cases gradually increased as the maximum and minimum humidity increased and vice versa.

**Figure 3:**
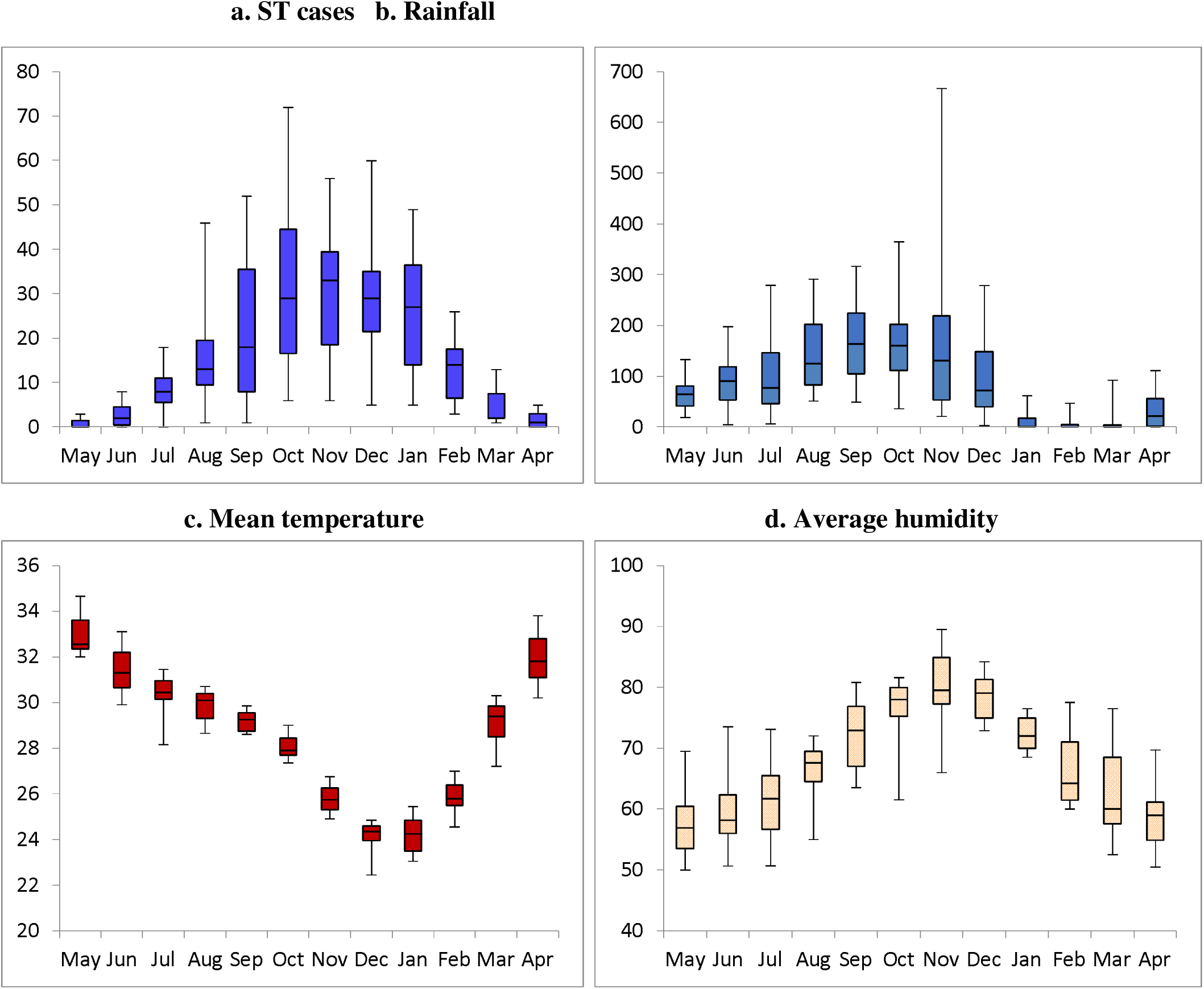
Consolidated Month-wise distribution of ST cases, rainfall, mean temperature and average humidity distribution of all 15 years.

Over the 15 years observed, there is a gradual increase of ST cases from the month of August to February. The ST cases were at peak from September to January and least number of cases were reported from March to July.

In Vellore, the average temperature during the cooler months (August to February) is 26.8°C (Range 22.9 – 31.9°C), the average humidity is 73.2% (Range 63.3 – 83.1%) with a median rainfall of 82.9 mm (IQR: 19.4 mm, 168.5 mm) during this period (Refer supplementary table No.1). The mean temperature in October and November (when scrub typhus cases peak) is 26.9°C (Range 22.6°C – 31.3°C), the average humidity is 78.4% (Range 71.6 – 85.1%) with a median rainfall of 140.5 mm (IQR: 70.9mm, 206.3mm) (Refer supplementary table No. 1).

The average temperature during the summer month (March – July) is 31.2°C (Range 24.9 - 37.4°C), the average humidity is 60.1% (Range 48.2 – 71.9%) with a median rainfall of 50.7mm (IQR: 5.7 mm, 97.7 mm) (Refer supplementary table No. 1).

**Table 3:** Month wise correlation between Scrub typhus cases and climate variables.

| Lag months | Rainfall |  | Mean temperature |  | Average humidity |  |
| --- | --- | --- | --- | --- | --- | --- |
|  | r | 95% CI | r | 95% CI | r | 95% CI |
| L_0 | 0.186* | 0.047, 0.322 | -0.698** | -0.773, -0.608 | 0.733** | 0.663, 0.790 |
| L_1 | 0.586** | 0.492, 0.656 | -0.452** | -0.555, -0.326 | 0.700** | 0.626, 0.755 |
| L_2 | 0.751** | 0.678, 0.809 | -0.037 | -0.167, 0.100 | 0.424** | 0.306, 0.527 |
| L_3 | 0.628* | 0.522, 0.717 | 0.383** | 0.269, 0.499 | 0.031 | -0.109, 0.164 |
| L_4 | 0.363** | 0.237, 0.492 | 0.656** | 0.557, 0.739 | -0.343** | -0.449, -0.225 |
| L_5 | 0.051 | -0.089, 0.193 | 0.732** | 0.651, 0.792 | -0.595** | -0.673, -0.490 |

\*\*Spearman's Correlation is significant at the 0.01 level (2-tailed).
\*Spearman's Correlation is significant at the 0.05 level (2-tailed).

For better understanding of the association between scrub typhus and climatic condition, Spearman’s correlation was done to find out the association between the time taken for a spurt in ST cases (lag time) and rainfall, temperature and humidity for up to a lag of 5 months. As observed earlier in the month-wise trend for 15 years, rainfall had the maximum positive correlation of 0.751 (95% CI: 0.678, 0.809) with a lag of two months and it is statistically significant (p <0.001). This positive association, however, reduced in the third and fourth month and became insignificant at the lag time of five months. Mean temperature showed a negative correlation of −0.698initially and reduced in lag of months and became insignificant in three lag months. However, it showed a positive correlation from 4 lag months.

Average maximum humidity showed a positive correlation with no lag and it continued till 2 lag months. Three lag months showed an insignificant correlation with four and five lag months showing a negative correlation.

**Table 4:** Spearman correlation coefficient (‘r’) matrix of meteorological variables.

|  | **Rainfall_2monthslag(mm)** | **Mean temperature (°C)** | **Mean relative humidity (%)** |
| --- | --- | --- | --- |
| Rainfall_2monthslag (mm) | 1 | - | - |
| Mean temperature (°C) | -0.612** | 1 | - |
| Mean relative humidity (%) | 0.618** | -0.739** | 1 |

All the p values are significant at 0.001 level

The above table describes the correlation coefficient matrix (Spearman) of the meteorological factors such as rainfall, mean temperature and mean relative humidity. Since the rainfall with 2 months lag has the highest correlation, we considered that for computing the Spearman correlation coefficient matrix and negative binomial regression. Whereas for the mean temperature and mean relative humidity, the same month reading with no lag was used. Mean temperature and mean relative humidity seem to be strongly correlated (−0.739) with each other.

**Table 5:**
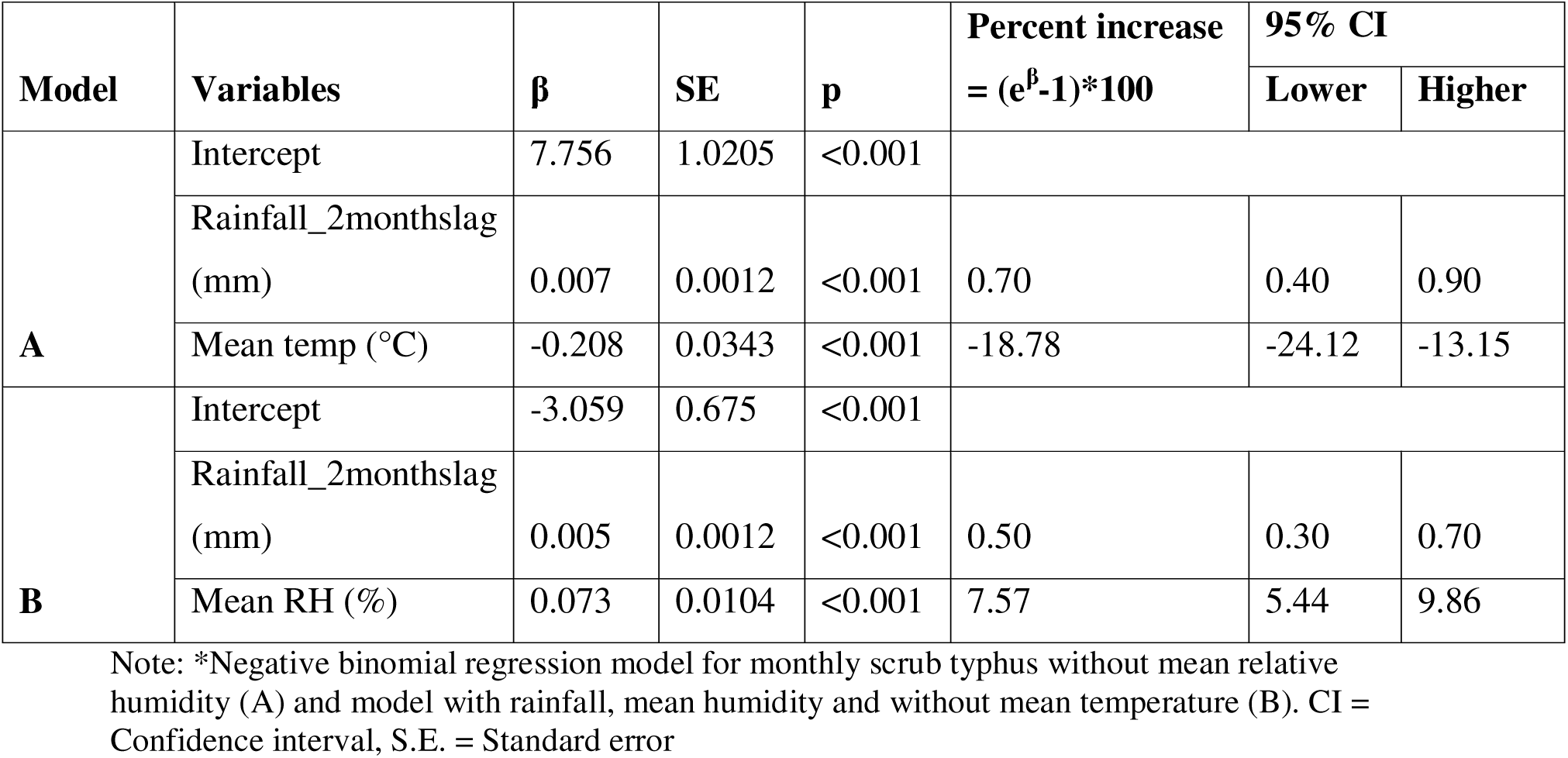
Negative Binomial regression analysis of climatic factors associated with scrub typhus in Vellore from May 2005 to April 2020.

To address the multicollinearity issues between mean temperatures and mean relative humidity in regression analysis, two different models (A & B) were developed using negative binomial regression for exploring the relation between mean temperature and mean relative humidity among ST cases. Model A was computed using the independent variables like rainfall_2monthslag and mean temperature. Model B with variables like rainfall_2months lag and mean humidity. For an increase of 1°C in mean temperature, the monthly ST case reduces by 18.78% (95% CI: −24.12, −13.15%). On the contrary, for 1 percent increase in mean relative humidity, there is an increase of 7.57% (95% CI: 5.44, 9.86%) of monthly ST cases. Similarly, for an increase of 1mm of rainfall contributes to increase of 0.50 to 0.70% of monthly ST cases after 2 months depending on the variables included in the analysis.

**Table 6:** Factors associated with Scrub Typhus.

| Variables |  | ST Positive | ST Negative | OR (95% CI) | p value | AOR (95% CI) | p value |
| --- | --- | --- | --- | --- | --- | --- | --- |
| Gender | Female | 1601<br>(29.5%) | 3832<br>(70.5%) | 1.55 (1.42 – 1.69) | <0.001 | 1.16 (1.05 – 1.30) | 0.006 |
|  | Male | 1183<br>(21.2%) | 4385<br>(78.8%) |  |  |  |  |
| Age category (years) | 0 – 20 (Ref) | 322<br>(15.8%) | 1714<br>(84.2%) | Reference |  |  |  |
|  | 21 – 30 | 399<br>(18.6%) | 1745<br>(81.4%) | 1.22 (1.04 – 1.43) | 0.017 | 0.67 (0.55 – 0.80) | <0.001 |
|  | 31 – 40 | 445<br>(26.2%) | 1254<br>(73.8%) | 1.89 (1.61 – 2.22) | <0.001 | 0.99 (0.82 – 1.20) | 0.911 |
|  | 41 – 50 | 499<br>(30.4%) | 1144<br>(69.6%) | 2.32 (1.98 – 2.72) | <0.001 | 1.13 (0.93 – 1.37) | 0.224 |
|  | 51 – 60 | 510<br>(32.1%) | 1078<br>(67.9%) | 2.52 (2.15 – 2.95) | <0.001 | 1.23 (1.02 – 1.50) | 0.034 |
|  | >60 | 609<br>(32.2%) | 1282<br>(67.8%) | 2.53 (2.17 – 2.95) | <0.001 | 1.37 (1.13 – 1.65) | 0.001 |
| Occupation | Farmers, daily wages & house wife | 1580<br>(33.8%) | 3100<br>(66.2%) | 2.74 (2.47 – 3.04) | <0.001 | 2.43 (2.13 – 2.77) | <0.001 |
|  | Others | 665<br>(15.7%) | 3577<br>(84.3%) |  |  |  |  |
| OP/IP | In-patient | 1871<br>(27.1%) | 5031<br>(72.9%) | 1.42 (1.30 – 1.56) | <0.001 | 1.11 (0.99-1.24) | 0.058 |
|  | Out-patient | 834<br>(20.7%) | 3186<br>(79.3%) |  |  |  |  |

### Univariate & Multivariate analysis

The association of ST with the risk factors were analyzed using chi-square and reported with 95% CI of Odds Ratio (OR). In univariate analysis, females, age group of >20 years, farmers, daily wage workers and house wives had the higher odds to be positive for scrub typhus when compared to males, age group 0 - 20 years and other occupations. In-patients also had a higher risk of being ST positive when compared to patients tested as out-patient. However, in multivariate analysis, females, age group of 51 – 60 years and >60 years, farmers, daily wage workers and house wives were at higher odds for being ST positive and the age group of 21 – 30 years showed a protective effect against ST when compared with 0 – 20 years of age.

## Discussion

In Vellore, scrub typhus is the commonest cause of Acute Undifferentiated Febrile Illness (AUFI), which is a group of acute fevers presenting without localizing signs (25).Scrub typhus (ST) is a vector borne disease (26), and its prevalence is related to various factors like vector abundance (27), climatic factors (28) and certain exposures like farming and owning domestic animals (29), outdoor activities (30) and sanitation (31). In this study, we wanted to assess how meteorological factors like temperature, rainfall and humidity and socio-economic factors impact the prevalence of scrub typhus using a 15 year dataset from a tertiary care hospital. The literature suggests that climatic factors like rainfall, temperature and humidity seem to have a positive impact on the occurrence of Scrub Typhus (24,32–34).

In this study, females were found to have a higher prevalence of scrub typhus (57.5%) than males and it is almost similar to the reported proportion of 58.7% in Vellore (35) and 51.46% in Guangzhou, Southern China (36). But a surveillance and case-control study done in Vietnam in 2021 reported less proportion of females being positive for scrub typhus (38.6%) (37). In multivariate analysis, females had a higher chance of being positive for scrub typhus. This is in agreement with previous studies from Tamil Nadu (38–40).

The prevalence of scrub typhus in this retrospective data set shows a steady upward trend from the third decade of life to a maximum prevalence of 21.1% among those older than 60 years. Similar age trend has been reported for scrub typhus in prospective sero-prevalence studies done in Northern Tamil Nadu (38,39) and in Bhutan (41). Same inferences have been obtained when retrospective data was analyzed in mainland China (40), Taiwan (32) and in Vientiane City, Lao (42). Other studies in Jharkhand (43) and North Bengal (12) have reported scrub typhus to be common in 1 to 20 years but the number of cases observed were less than 100.

The majority of scrub typhus positives in our study came from the in-patient department, which in our opinion is due to a reference center bias, as more sick patients who need hospitalization are referred to a tertiary care facility (25). The occupations at high risk in this study are farmers, daily wage laborers and house wives which is quite similar to the study from Guangzhou, Southern China (36). Agricultural work has been identified as a risk factor for acquiring scrub typhus in India from Tamil Nadu (34 & 37, 43) (38), Darjeeling (44). Similar findings have been reported by researchers in Bhutan (41), China (40), Nepal (45), Thailand (46), South Korea (47) and Japan (48).

A clear seasonality of scrub typhus occurrence in Vellore district was noticed with maximum cases occurring from August to February with peak in October and November. The above finding reiterates the observation that scrub typhus cases are maximum in the cooler months of the year in Vellore as reported by Mathai et al in 2003(19) and Abhilash et al in 2016 (25). The primary peak of ST was observed in Jiangxi Province, China in September and October, and a smaller peak from June to August (49). Another study from Guangzhou, Southern China in 2021 reported high number of ST cases during the months of May to October with a peak in June (36), from June to September in Vientiane, Laos (50) and June to November in Taiwan (32). In North-East India cases of scrub typhus occur in July to November in Manipur (51), July to October in Sikkim (52) and September to November in Darjeeling (44).

Rainfall and Scrub Typhus had a positive correlation of 0.751 (p <0.001) with a lag of two months. For an increase of 1mm of rainfall, 0.50 to 0.70% of ST cases rise after two months in our study which is higher than the reported 0.05% to 0.10% ST cases with 1mm rise in rainfall in Guangzhou, Southern China (24). On the contrary, there was an increase of 5.23% of ST cases for every 1mm rise in the rainfall in Taiwan (32). A prospective study conducted in a hospital in Uttar Pradesh reported a positive association of scrub typhus and rainfall as the majority of the cases were observed in the post-monsoon season, but the rainfall associated increase is not mentioned(53). The same trend was observed in the study done in Laos (50) and in Vellore (35) though the associated increase was not documented. The current study documents the associated increase of scrub typhus with variation in meteorological factors.

Mean temperature had a negative correlation with the ST cases in our study which is contradicted by the spatio-temporal analysis in Laos, where the maximum temperature had a positive impact on ST cases (50). In our study, the maximum number of cases occurred in the months with a mean temperature is 26.8°C (implying that lower temperature is more favorable for ST to occur in Vellore, as the summer mean temperature is 31°C). This is in agreement with the findings reported from previous studies done in this region (19,54). In contrast, a multi-centric study in Southern Taiwan reported a positive correlation of ST with increase in temperature. However, the maximum number of cases were reported in summer months where the average temperature ranged from 25 – 30°C (55). A spatial risk analysis done in Jiangsu Province, China reported a suitable temperature during March - November (Summer type) for the ST ranging from 31.7 – 32.8°C (56), which is slightly higher than what we have observed in this study. In spite of having a negative correlation of temperature in our study, the temperature which is favorable for ST is almost similar with the other studies done elsewhere in the tsutsugamushi triangle. In Southern India, scrub typhus occurs in cooler months (19), whereas in China it occurs mainly in the summer months (57). However, the suitable temperature for the prevalence of scrub typhus is almost similar in all the reported countries leaving the impression that the temperature is more important than comparing the seasons.

For an increase of 1°C of temperature, the monthly ST case reduces by 18.78% (95% CI: - 24.12, −13.15%) in our study. At initial scrutiny it looks like contradicting to the reported increase of 14.98% of monthly ST cases in Guangzhou, Southern China from 2006 – 2012 (24). A retrospective study done by analyzing the 15 years scrub typhus data in Korea reported that for an increase of 1°C in temperature, there is an increase of 38 ST cases during the month of May or June (33) and an increase of 3.279 cases for every 1 °C temperature increase in Taiwan (32). This discrepancy of the result is because of the average temperature during the cooler month’s (August to February) ranges from 7 to 29°C in Guangzhou, China (https://www.timeanddate.com/weather/@8513624/climate), −1 to 26°C in Jeollabuk-do, Korea (https://www.timeanddate.com/weather/@1845789/climate), and in Vellore it is 22.9 – 31.9°C.This clearly demonstrates that extreme temperatures are not suitable for the spread of scrub typhus. The suitable temperature range for the trombiculid mites to lay eggs is 20 to 30°C (58) and therefore based on temperature, the data from tropical India and temperate East-Asian countries is in agreement.

Humidity, on the other hand had a positive influence on ST cases and the same was observed in Laos (50), in Guangzhou, Southern China (21,36) and in Thailand by Bhopdhornangkul et al (34). For 1 percent increase in mean relative humidity, there was an increase of 7.57% (95% CI: 5.44, 9.86%) of monthly ST cases in our study. This is low when compared to the increase of monthly ST cases by 33% for an increase of 1% of maximum relative humidity in that particular month in Thailand (34).

## Conclusion

This study explains the influence of the climate on Scrub Typhus in Vellore, Tamil Nadu. Rainfall is found to be a major influencing factor in the occurrence of Scrub Typhus cases with a lag of two months in all fifteen years. Temperature had a negative correlation as the number of scrub typhus cases diagnosed increased in the cooler months and very minimal or nil cases were observed during the hot summer months of Vellore. Humidity on the other hand proved to be positively correlated with the Scrub Typhus and more number of cases was observed in months with mean relative humidity from 55 to 89%. This study suggests the healthcare professionals to be prepared for the early diagnosis and treatment of Scrub Typhus during the rainy season and cooler months with increased humidity. Scrub Typhus prevention measures like health education for high-risk populations such as farmers, farming-related occupations and housewives can be provided.

## Supporting information

Suppl Data - Climate & Scrub typhus Vellore.docx.

## Data Availability

All data produced in the present study are available upon reasonable request to the authors

## Ethics approval

This study was conducted after approval by the Institutional Review Board (IRB) and Ethics committee vide IRB Min no. 9866 dated 20^th^ January 2016.

## Acknowledgements

None

