## Supplementary material for "Climate influences scrub typhus occurrence in Vellore, Tamil Nadu, India: Analysis of a 15 year dataset": Suppl Data - Climate & Scrub typhus Vellore.docx.

**Supplementary Data**

**Table 1: Summary of monthly scrub typhus confirmed cases and weather conditions in Vellore, May 2005 - April 2020**

| **Monthly Meteorological factors** | **Mean** | **SD** | **Min** | **25%** | **50%** | **75%** | **Max** |
| --- | --- | --- | --- | --- | --- | --- | --- |
| ST cases | 15.47 | 16.08 | 0 | 2 | 9.5 | 23.75 | 72 |
| Rainfall in mm | 88.81 | 92.92 | 0 | 9.67 | 64 | 135.2 | 667.2 |
| Mean temperature | 28.62 | 2.94 | 22.45 | 25.80 | 29.00 | 30.85 | 34.85 |
| Mean Humidity | 67.72 | 9.15 | 50 | 60.00 | 68.15 | 75.25 | 89.50 |

**Table 2: Consolidated monthly scrub typhus cases and weather conditions in Vellore**

| **Month** | **Scrub typhus cases** | | **Rainfall in mm** | | **Temperature** | | **Humidity** | |
| --- | --- | --- | --- | --- | --- | --- | --- | --- |
|  | **Mean ± SD** | **Median (IQR)** | **Median** | **IQR** | **Mean** | **SD** | **Mean** | **SD** |
| January | 26.53 **±** 13.77 | 27 (13, 38) | 0.70 | 0, 17.40 | 24.43 | 1.34 | 72.22 | 2.59 |
| February | 12.33 **±** 6.91 | 14 (5, 18) | 0 | 0, 4.80 | 25.96 | 0.72 | 66.15 | 5.76 |
| March | 4.47 **±** 3.66 | 2 (2, 8) | 0 | 0, 4 | 29.16 | 0.88 | 62.62 | 6.66 |
| April | 1.53 **±** 1.69 | 1 (0, 3) | 21.60 | 1.90, 56.10 | 31.92 | 0.97 | 59.08 | 5.05 |
| May | 0.73 **±** 1.03 | 0 (0, 2) | 64 | 41.40, 81 | 32.93 | 0.86 | 57.74 | 4.96 |
| June | 2.67 **±** 2.64 | 2 (0, 5) | 90.30 | 53, 118.30 | 31.42 | 0.97 | 59.69 | 6.05 |
| July | 8.27 **±** 5.13 | 8 (5, 13) | 77 | 45.80, 146.40 | 30.46 | 0.93 | 61.24 | 6.04 |
| August | 14.53 **±** 11.14 | 13 (9, 21) | 125.10 | 82.90, 202.20 | 30.10 | 0.72 | 66.47 | 4.37 |
| September | 22.47 **±** 17.33 | 18 (7, 36) | 163.30 | 104.20, 224.30 | 29.23 | 0.49 | 72.26 | 5.01 |
| October | 32.80 **±** 20.02 | 29 (13, 50) | 160.40 | 111.60, 202 | 28.06 | 0.61 | 76.64 | 5.07 |
| November | 30.13 **±** 15.19 | 33 (18, 40) | 130.20 | 53.30, 219 | 25.79 | 0.56 | 80.06 | 5.60 |
| December | 29.13 **±** 14.53 | 29 (20, 36) | 72.40 | 39.50, 148.60 | 24.20 | 0.70 | 78.47 | 3.70 |

**Table 3: Year-wise distance from scrub patient’s house to CMC, Vellore in Km from May 2005 – April 2020**

| **S.no** | **Years** | **Distance from scrub patient’s home to CMC, Vellore in km** | | | | |
| --- | --- | --- | --- | --- | --- | --- |
|  |  | **Mean** | **SD** | **Median** | **25%** | **75%** |
| 1 | May - 05 - Apr 06 | 18.15 | 19.79 | 13.37 | 4.12 | 24.23 |
| 2 | May - 06 - Apr 07 | 20.94 | 21.34 | 14.36 | 4.55 | 28.63 |
| 3 | May 07 - Apr 08 | 21.51 | 20.93 | 15.64 | 5.20 | 30.97 |
| 4 | May 08 - Apr 09 | 25.23 | 22.87 | 21.39 | 5.53 | 30.91 |
| 5 | May 09 - Apr 10 | 26.36 | 22.68 | 22.39 | 7.30 | 34.09 |
| 6 | May 10 - Apr 11 | 23.57 | 23.41 | 18.35 | 4.95 | 30.97 |
| 7 | May 11 - Apr 12 | 31.40 | 26.55 | 22.21 | 8.18 | 51.04 |
| 8 | May 12 - Apr 13 | 24.42 | 22.51 | 18.96 | 5.53 | 34.09 |
| 9 | May 13 - Apr 14 | 23.53 | 21.76 | 17.72 | 5.53 | 31.17 |
| 10 | May 14 - Apr 15 | 28.93 | 23.38 | 24.23 | 11.16 | 35.92 |
| 11 | May 15 - Apr 16 | 26.28 | 24.14 | 20.88 | 5.56 | 33.94 |
| 12 | May 16 - Apr 17 | 25.98 | 21.44 | 20.70 | 11.55 | 33.69 |
| 13 | May 17 - Apr 18 | 28.20 | 22.91 | 21.50 | 11.58 | 36.21 |
| 14 | May 18 - Apr 19 | 20.91 | 18.74 | 16.08 | 6.55 | 29.14 |
| 15 | May 19 - Apr 20 | 18.09 | 17.54 | 12.96 | 5.68 | 24.61 |

**Figure 1: Scrub typhus tested, positive and positivity rate from May 2005 to April 2020**

|  | Tested | Positive | ST Positivity Rate |
| --- | --- | --- | --- |
| May 05 - Apr 06 | 128 | 32 | 25% |
| May 06 - Apr 07 | 336 | 87 | 25.8929% |
| May 07 - Apr 08 | 585 | 232 | 39.6581% |
| May 08 - Apr 09 | 563 | 182 | 32.3268% |
| May 09 - Apr 10 | 523 | 127 | 24.283% |
| May 10 - Apr 11 | 580 | 216 | 37.2414% |
| May 11 - Apr 12 | 705 | 305 | 43.2624% |
| May 12 - Apr 13 | 1218 | 298 | 24.4663% |
| May 13 - Apr 14 | 696 | 283 | 40.6609% |
| May 14 - Apr 15 | 475 | 121 | 25.4737% |
| May 15 - Apr 16 | 620 | 180 | 29.0323% |
| May 16 - Apr 17 | 687 | 159 | 23.1441% |
| May 17 - Apr 18 | 1029 | 200 | 19.4363% |
| May 18 - Apr 19 | 805 | 161 | 20% |
| May 19 - Apr 20 | 2051 | 201 | 9.8001% |

**Figure 2: Year-wise gender distribution of ST cases**

|  | Male | Female |
| --- | --- | --- |
| May 05 - Apr 06 | 16 | 16 |
| May 06 - Apr 07 | 38 | 49 |
| May 07 - Apr 08 | 104 | 128 |
| May 08 - Apr 09 | 80 | 102 |
| May 09 - Apr 10 | 64 | 63 |
| May 10 - Apr 11 | 101 | 115 |
| May 11 - Apr 12 | 120 | 185 |
| May 12 - Apr 13 | 130 | 168 |
| May 13 - Apr 14 | 109 | 174 |
| May 14 - Apr 15 | 48 | 73 |
| May 15 - Apr 16 | 77 | 103 |
| May 16 - Apr 17 | 63 | 96 |
| May 17 - Apr 18 | 89 | 111 |
| May 18 - Apr 19 | 60 | 101 |
| May 19 - Apr 20 | 84 | 117 |

**Figure 3: Occupation-wise ST cases from 2005 – 2020**

|  | Negative | Positive | ST - Positivity rate |
| --- | --- | --- | --- |
| Preschoolers | 372 | 79 | 21.2366% |
| Student | 1428 | 311 | 21.7787% |
| House wife | 1875 | 891 | 47.52% |
| Daily wages | 921 | 517 | 56.1346% |
| Farmer | 304 | 172 | 56.5789% |
| Healthcare professionals | 287 | 25 | 8.7108% |
| Retired | 206 | 56 | 27.1845% |
| Others | 1284 | 194 | 15.109% |

**Figure 4: Month-wise ST case distribution in 15 years**

|  | 2005 - 06 | 2006 - 07 | 2007 - 08 | 2008 - 09 | 2009 - 10 | 2010 - 11 | 2011 - 12 | 2012 - 13 | 2013 - 14 | 2014 - 15 | 2015 - 16 | 2016 - 17 | 2017 - 18 | 2018 - 19 | 2019 - 20 |
| --- | --- | --- | --- | --- | --- | --- | --- | --- | --- | --- | --- | --- | --- | --- | --- |
| May | 0 | 0 | 0 | 2 | 0 | 0 | 2 | 1 | 2 | 0 | 1 | 0 | 0 | 3 | 0 |
| June | 0 | 1 | 4 | 4 | 2 | 2 | 8 | 0 | 5 | 1 | 6 | 0 | 1 | 6 | 0 |
| July | 0 | 2 | 8 | 8 | 9 | 14 | 6 | 7 | 13 | 5 | 16 | 18 | 6 | 9 | 3 |
| August | 2 | 1 | 18 | 10 | 4 | 22 | 13 | 21 | 24 | 10 | 13 | 46 | 10 | 9 | 15 |
| September | 1 | 1 | 41 | 35 | 3 | 20 | 50 | 17 | 52 | 9 | 16 | 18 | 36 | 7 | 31 |
| October | 6 | 11 | 29 | 27 | 36 | 33 | 50 | 72 | 64 | 11 | 27 | 13 | 54 | 20 | 39 |
| November | 6 | 19 | 19 | 17 | 12 | 40 | 47 | 52 | 56 | 33 | 18 | 22 | 36 | 39 | 36 |
| December | 7 | 29 | 34 | 42 | 20 | 29 | 47 | 60 | 28 | 23 | 14 | 5 | 36 | 29 | 34 |
| January | 5 | 13 | 42 | 22 | 11 | 35 | 46 | 38 | 27 | 22 | 49 | 15 | 12 | 34 | 27 |
| February | 3 | 8 | 19 | 14 | 20 | 10 | 26 | 18 | 9 | 5 | 14 | 17 | 5 | 3 | 14 |
| March | 2 | 2 | 13 | 1 | 8 | 8 | 7 | 9 | 2 | 2 | 2 | 5 | 3 | 2 | 1 |
| April | 0 | 0 | 5 | 0 | 2 | 3 | 3 | 3 | 1 | 0 | 4 | 0 | 1 | 0 | 1 |

**Figure 5: Consolidated Month-wise ST case distribution of all 15 years vs Rainfall, Temperature and Humidity**

|  | ST Cases | Mean Max Temp (°F) | Mean min Temp (°F) | Average Rainfall (mm) | RH 08:30 (%) | RH 17:30 (%) |
| --- | --- | --- | --- | --- | --- | --- |
| May | 11 | 103.28 | 79.34 | 65.2 | 67.6 | 47.9 |
| June | 40 | 98.6 | 78.44 | 92.2 | 67.6 | 51.8 |
| July | 124 | 96.26 | 77.36 | 106.2 | 69.1 | 53.4 |
| August | 218 | 94.64 | 76.64 | 147 | 74 | 58.9 |
| September | 337 | 93.2 | 76.1 | 168.3 | 78.4 | 66.1 |
| October | 492 | 90.5 | 74.48 | 162.3 | 83.5 | 69.8 |
| November | 452 | 86 | 70.88 | 166.7 | 86.6 | 73.5 |
| December | 437 | 83.84 | 67.28 | 91.4 | 88.2 | 68.8 |
| January | 398 | 86.9 | 65.12 | 13.4 | 87.2 | 57.3 |
| February | 185 | 91.04 | 66.38 | 8.1 | 83.9 | 48.4 |
| March | 67 | 96.98 | 72.14 | 12.5 | 80.4 | 44.8 |
| April | 23 | 101.66 | 77.18 | 32.4 | 74.8 | 43.4 |
